# A novel hospital-at-home model for patients with COVID-19 built by a team of local primary care clinics and clinical outcomes: A multi-center retrospective cohort study

**DOI:** 10.1101/2022.10.28.22281588

**Authors:** Yasushi Tsujimoto, Masanori Kobayashi, Tomohisa Oku, Takahisa Ogawa, Shinichi Yamadera, Masako Tsukamoto, Noriya Matsuda, Morikazu Nishihira, Yu Terauchi, Takahiro Tanaka, Yoshitaka Kawabata, Yuki Miyamoto, Yoshiki Morikami, KISA2-Tai Osaka

## Abstract

**Background:** Hospital-at-home (HaH) care has been proposed as an alternative to inpatient care for patients with COVID-19. Previous reports were hospital-led and involved patients triaged at the hospitals. To reduce the burden on hospitals, we constructed a novel HaH care model organised by a team of local primary care clinics.

**Methods:** We conducted a multi-center retrospective cohort study of the COVID-19 patients who received our HaH care from Jan 1^st^ to Mar 31^st^, 2022. Patients who were not able to be triaged for the need for hospitalization by the Health Center solely responsible for the management of COVID-19 patients in Osaka City were included. The primary outcome was receiving medical care beyond the HaH care defined as a composite outcome of any medical consultation, hospitalization, or death within 30 days from the initial treatment.

**Results:** Of 382 eligible patients, 34 (9%) were triaged for hospitalization immediately after the initial visit. Of the remaining 348 patients followed up, 37 (11%) developed the primary outcome, while none died. Obesity, fever, and gastrointestinal symptoms at baseline were independently associated with an increased risk of needing medical care beyond the HaH care. A further 129 (37%) patients were managed online alone without home visit, and 170 (50%) required only one home visit in addition to online treatment.

**Conclusions:** The HaH care model with a team of primary care clinics was able to triage patients with COVID-19 who needed immediate hospitalization without involving hospitals, and treated most of the remaining patients at home.

## Introduction

The global pandemic of coronavirus disease (COVID-19) caused by a novel coronavirus (severe acute respiratory coronavirus 2, SARS-CoV-2) first identified in Wuhan in 2019 has spread rapidly worldwide (1). As SARS-CoV-2 spreads, health care systems continue to face the threat of collapse from the rapidly growing number of COVID-19 patients, some of whom, primarily the elderly and those with comorbidities such as obesity, diabetes, and serious cardiac disease, require hospitalization (2, 3). A surge in hospital admissions has increased the burden on medical resources such as beds for patients with infectious diseases, intensive care units, ventilators, personal protective equipment, and medical staff. This increased burden threatens the quality of healthcare not only for both COVID-19 and other patients (4, 5).

Hospital at home (HaH), a model of care in which acute care is provided in the patient’s home at a level previously provided in a hospital, has been proposed as a potentially useful alternative to inpatient care (6). Regarding the utility of HaH for COVID-19 patients, Atrium Health, the largest integrated healthcare service in the United States, reported a study of a virtual hospital (7); thereafter, all similar efforts were hospital-led (8-11). Patients were diagnosed with COVID-19 at the hospitals and were triaged to be hospitalized or not. If the patients did not require hospitalization or recovered after hospitalization, HaH care was provided by the hospital team. However, it is difficult for a hospital to secure the personnel to provide a HaH care only for a period of the surge of COVID-19 patients. Thus, the development of a HaH system that would reduce the burden on hospitals during this period is important.

To reduce the burden on hospitals, it is crucial to triage the patients who require immediate hospitalization and to provide care for COVID-19 patients in the primary care setting rather than in hospital. We previously reported the usefulness of HaH care without hospital visits for elderly COVID-19 patients in Kyoto city (12). This provision of HaH care in a primary care clinic that combines the functions of triage and inpatient care is unique worldwide. However, the model was based on one clinic, and this may be difficult to apply to other settings, or to cover large area. We therefore formed a team of ten different clinics to cover the entirety of Osaka city, one of Japan’s largest cities, with the highest number of deaths and mortality rate (13). This study aimed to describe the detailed characteristics, treatments, and clinical outcomes of patients who received HaH care by a team of general clinics.

## Methods

### Study design and setting

We conducted a multi-center retrospective cohort study from Jan 1^st^ to Mar 31^st^, 2022 when the BA.2 lineage was predominant (14). We followed the STROBE statement for reporting of cohort studies (15), and registered the pre-specified protocol in the University Hospital Medical Information Network (UMIN000047837). The Strengthening the Reporting of Observational Studies in Epidemiology (STROBE) checklist (16) and the difference in the pre-specified protocol and publication was described in the Supplementary Table 1 and Supplementary Table 2. As this is a retrospective study of routinely obtained data, written informed consent was waived. An opportunity for refusal to participate in research was guaranteed by an opt-out manner. The study was approved by the Ethics Committee of Japan Primary Care Association (ethics approval number: R2628).

### Setting

We rapidly developed a hospital at home model with a team of 10 different clinics located in Osaka city (The “Kansai Intensive Area Care Unit for SARS-CoV-2 (KISA2-Tai) Osaka). To cover the entirety of the Osaka city area, one of the 10 clinics was responsible for the initial treatment on a rotating basis, with follow-up care provided by the clinic closest to the patient’s home. During the study period, Osaka City recommended patients with risk factors for developing severe disease to be hospitalized regardless of their severity. Further, patients diagnosed with COVID-19 were required to be quarantined for 10 days. The Osaka City Health Center consolidated the information on all the cases of COVID-19 diagnosed at each medical institution in Osaka city and triaged the need for hospitalization. The Health Center submitted a request for medical treatment to the KISA2-Tai Osaka i) if they could not triage the patients who required immediate hospitalization through a phone call with the patient, ii) if hospitalization was difficult due to a shortage of hospital beds or due to patients’ refusal, or iii) if no primary care clinic near their homes could treat their emergencies.

### Participants

All patients diagnosed as COVID-19 and treated by the KISA2-Tai Osaka upon the request from the Osaka City Health Center during the study period were eligible. The diagnosis of COVID-19 was based on nucleic acid amplification tests, antigen tests, or clinical investigation. Patients who disagreed to participate this study were excluded.

### Data sources and variables

To help clinics collaborate with each other, we used a uniform medical interview sheet investigating patient information. The information was routinely stored in a secured database. In addition to the general patient characteristics, the following demographics were sought from the database and electrical record: day of the week of the initial treatment (Monday to Thursday or Friday to Sunday), ability to walk, vaccination history (less than twice or not), persons living with the patients (alone, children younger than 18 years old, or other) ability to procure food on their own, health insurance (public assistance or not), and whether the patients had a family doctor. Since the Kisa2-Tai Osaka used a uniform medical interview sheet to share patient information across different clinics, the information about the known risk factors for severe COVID-19, past medical history and symptoms were available. Smoking, body mass index, malignancy, chronic respiratory disease, chronic kidney disease, diabetes, dyslipidemia, post-transplant immunodeficiency, dementia, and pregnancy depression were included as variables for risk factors or past medical history. Peripheral oxygen saturation at the initial treatment, fever, headache, dyspnea, cough, sore throat, gastrointestinal symptoms, losing taste, fatigue, chest pain, and water intake (less than 500 ml or not) were included as symptom variables. We further calculated the straight distance between the clinic responsible for the initial treatment and the patient’s home using the latitude and longitude of the postal code. The electronic medical records at each clinic were used to extract information on the details of the treatment, including the number of doctor visits at home, number of online doctor visits, number of types of prescriptions, number of days of intravenous infusion, and whether corticosteroids, Remdesivir, Molnupiravir, or Sotrovimab were administered.

The primary outcome was receiving medical care beyond the HaH care, defined as a composite outcome of any medical consultation outside of the team, hospitalization, or death due to any cause within 30 days from the initial treatment. The secondary outcomes were each component of the primary outcome.

### Statistical analysis

The roles of the team were i) to triage patients who required immediate hospital-based medical examinations or interventions and ii) to provide at-home treatment to patients who did not need to be hospitalized during the study period. We therefore excluded the patients who were admitted immediately after the initial treatment (“ruled-in” patients), and those who were followed up by the team after the initial treatment (“ruled-out” patients) were included in the main analysis.

For the main analysis, we used the ruled-in patients and tabulated the baseline characteristics, symptoms, and details of treatment provided by the HaH team, stratified by receiving medical care beyond the team or not. We further describe the number and proportion of patients with the secondary outcomes. For patients who had the primary outcome, we narratively summarized the reason why they needed medical care beyond the HaH team. As an exploratory analysis, we used a mixed effect logistic regression model adjusted with a random effect for each clinic responsible for the initial treatment to identify potential factors associated with the primary outcome. In this model, we included the following pre-specified baseline covariates: age, sex, vaccination, living with young children, living alone, ability to walk, public assistance insurance, existence of family physicians, chronic respiratory disease, hypertension, arrhythmia, obesity, hyperlipidemia, diabetes mellitus, chronic kidney disease, depression, dementia, malignancy, peripheral oxygen saturation, fever, cough, dyspnea, gastrointestinal symptoms, water intake, headache, sore throat, losing taste, fatigue, and chest pain.

As this study was primarily descriptive in purpose, the sample size was not calculated. The *p*-value was two-sided, and values <0.05, were deemed statistically significant. All analyses were performed using Stata SE, version 14.2 (StataCorp, College Station, TX).

## Results

During the study period, 384 patients were admitted to the HaH care team for triage and treatment. As shown in Figure 1, two patients were excluded due to loss to follow-up and refusal to participate, leaving 382 patients included in this study. Thirty-four patients were hospitalized immediately after the initial triage by our team. The characteristics of the patients requiring immediate hospitalization are described in Supplementary Table 2.

**Figure 1.**
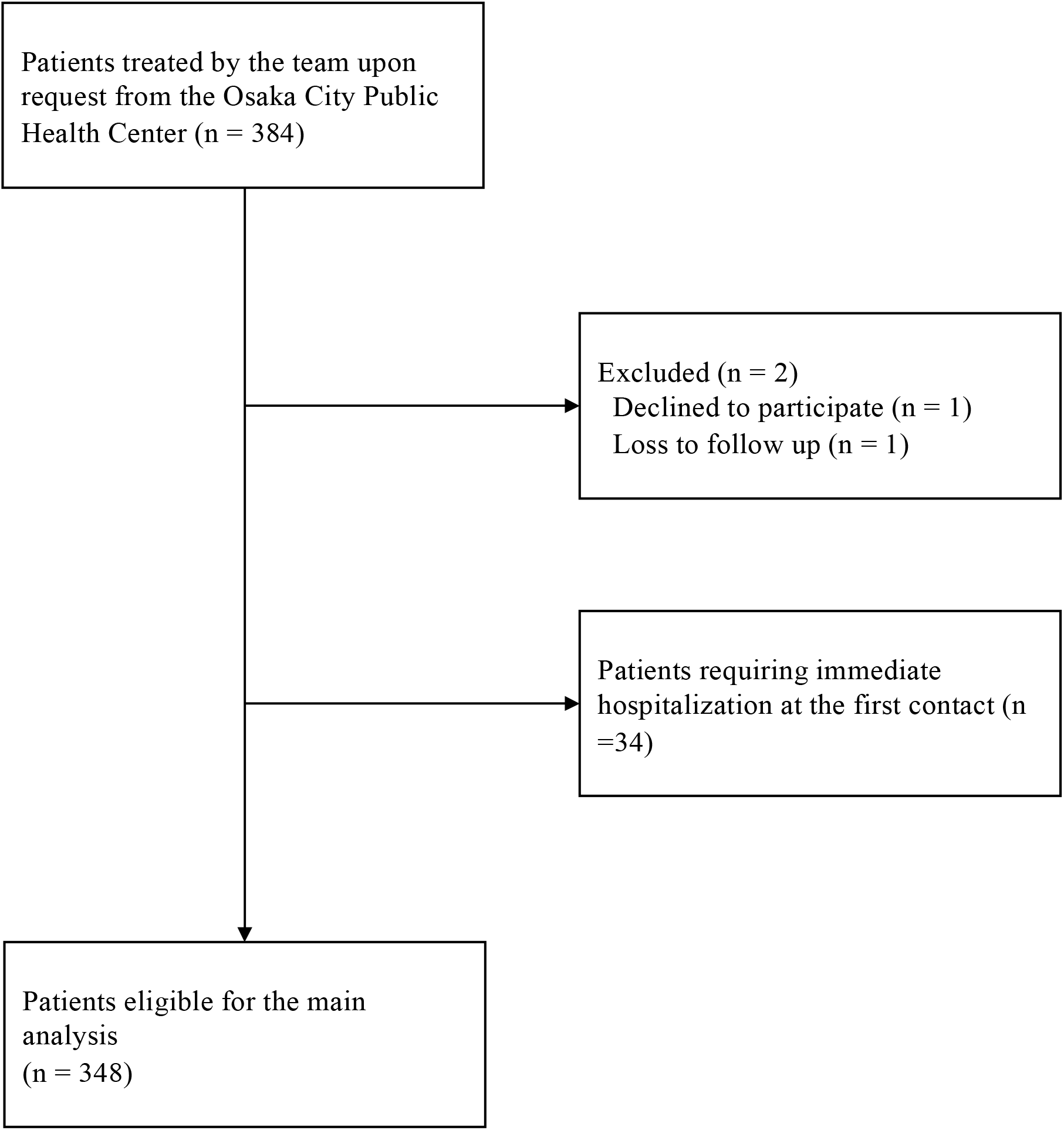
Patient flow diagram.

Figure 2 illustrates the number of doctor visit requests from the Health Center and hospital capacity for mild to moderate COVID-19 patients in Osaka. The number of requests rapidly increased in late January, and the hospital capacity in Osaka peaked on 14^th^ Feb. Table 1 describes the characteristics of 348 patients who were followed up by our team, stratified by receiving medical care beyond the team. The number of missing values in each variable is shown in Supplementary Table 4. A total of 37 patients (11%) required medical care beyond the HaH care, including 8 (2%) who required medical consultation outside of the team and 29 (8%) who were discharged alive after hospitalization. These patients tended to be older, needed major help to walk, with hypoxia and symptoms such as fever, cough, and difficulty drinking at baseline. There were no deaths due to any cause within 30 days from the initial treatment, either in the HaH or in the hospital. The reasons for the requirement of medical care beyond HaH care are summarized in Supplementary Table 5. Hypoxia was the most frequent reason for hospitalization, and the median (interquartile range) time from the onset to medical care beyond the HaH care was 11 (7 to 13) days. Of the 13 patients admitted for hypoxia, 9 had oxygen saturation greater than 94% at baseline.

**Table 1.** Baseline characteristics of the patients followed by the hospital-at-home care team.

| Characteristics | All (n =348) | Received medical care solely by the HaH care (n = 311) | Received medical care beyond the HaH care<sup>†</sup> (n = 37) | p-value* |
| --- | --- | --- | --- | --- |
| **General** |  |  |  |  |
| Age | 44.5 (30.5 to 66) | 42 (30 to 64) | 61 (48 to 73) | <0.001 |
| Male gender | 153 (44) | 133 (43) | 20 (54) | 0.19 |
| Days from onset | 6 (3 to 8) | 6 (3 to 8) | 6 (4 to 9) | 0.71 |
| Unvaccinated<sup>‡</sup> | 164 (51) | 142 (46) | 22 (56) | 0.26 |
| Ability to walk |  |  |  |  |
| Independent | 280 (88) | 258 (90) | 22 (60) | <0.001 |
| Minor help | 13 (4) | 11 (4) | 2 (5) |  |
| Major help | 21 (7) | 13 (5) | 8 (22) |  |
| Bedridden | 5 (2) | 4 (1) | 1 (3) |  |
| Living with children under 18 | 123 (41) | 119 (44) | 4 (11) | <0.001 |
| Living alone | 61 (19) | 51 (18) | 10 (27) | 0.089 |
| Public assistance insurance | 26 (8) | 24 (8) | 2 (5) | 0.61 |
| Having a family physician | 185 (64) | 159 (62) | 26 (70) | 0.16 |
| Food procurement<sup>§</sup> | 261 (89) | 229 (88) | 32 (87) | 0.12 |
| Weekend requests for HaH care | 86 (25) | 74 (24) | 12 (32) | 0.25 |
| Distance (km) | 5.7 (3.0 to 9.7)<br>(n = 197) | 5.9 (3.2 to 9.9)<br>(n = 170) | 5.1 (1.8 to 6.9)<br>(n = 27) | 0.059 |
| **Risk factors** |  |  |  |  |
| Chronic respiratory disease | 64 (18) | 59 (19) | 5 (14) | 0.42 |
| Hypertension | 76 (22) | 58 (19) | 18 (49) | <0.001 |
| Arrhythmia | 16 (5) | 14 (5) | 2 (5) | 0.80 |
| Obesity | 83 (24) | 67 (22) | 16 (43) | 0.003 |
| Hyperlipidemia | 36 (10) | 29 (9) | 7 (19) | 0.070 |
| Diabetes mellitus | 37 (11) | 27 (9) | 10 (27) | <0.001 |
| Chronic kidney disease | 14 (4) | 12 (4) | 2 (5) | 0.65 |
| Depression | 13 (4) | 12 (4) | 1 (3) | 0.73 |
| Dementia | 5 (1) | 3 (1) | 2 (5) | 0.032 |
| Malignancy | 34 (10) | 27 (9) | 7 (19) | 0.047 |
| Pregnant | 4 (1) | 4 (1) | 0 (0) | 0.49 |
| **Symptoms at the initial treatment** |  |  |  |  |
| Peripheral oxygen saturation<sup>**</sup> |  |  |  | 0.077 |
| 96% ≤ | 290 (83) | 264 (85) | 27 (70) |  |
| 93% to 96% | 33 (9) | 27 (9) | 6 (16) |  |
| 93% ≥ | 25 (7) | 20 (6) | 5 (14) |  |
| Fever | 140 (40) | 118 (38) | 22 (60) | 0.012 |
| Cough | 263 (80) | 241 (82) | 22 (60) | 0.003 |
| Dyspnea | 141 (43) | 123 (42) | 18 (49) | 0.44 |
| Gastrointestinal symptom | 124 (38) | 107 (37) | 17 (46) | 0.18 |
| Water intake of less than 500ml per day | 37 (11) | 30 (10) | 7 (22) | 0.089 |
| Headache | 147 (46) | 128 (45) | 19 (51) | 0.40 |
| Sore throat | 176 (56) | 159 (56) | 17 (46) | 0.38 |
| Losing taste | 74 (24) | 65 (24) | 9 (24) | 0.72 |
| Fatigue | 239 (74) | 210 (73) | 29 (78) | 0.31 |
| Chest pain | 85 (27) | 75 (27) | 10 (27) | 0.90 |

Note: Values in parentheses shows percentage or interquartile range. Percentage calculations used a denominator that excluded missing values. \* *p*-values based on the Fisher's exact test for continuous variables and the Pearson's chi-squared test for categorical variables. † A composite outcome of any medical consultation outside of the HaH care team, hospitalization, or death due to any cause within 30 days from the initial treatment. ‡ No vaccination or only first vaccination against Coronavirus disease § Living with children under 18 years old. § Inability to procure food without stockpiles as declared by the individual. || Requests for HaH care made from Friday to Sunday. ¶ Only patients who needed doctor visits at home for the initial treatment were included, and the straight-line distance from the clinic where the initial treatment was provided to the patient's home was calculated using latitude and longitude. \*\* Post-exertion oxygen saturation if applicable. Abbreviations: HaH, hospital at home

**Figure 2.**
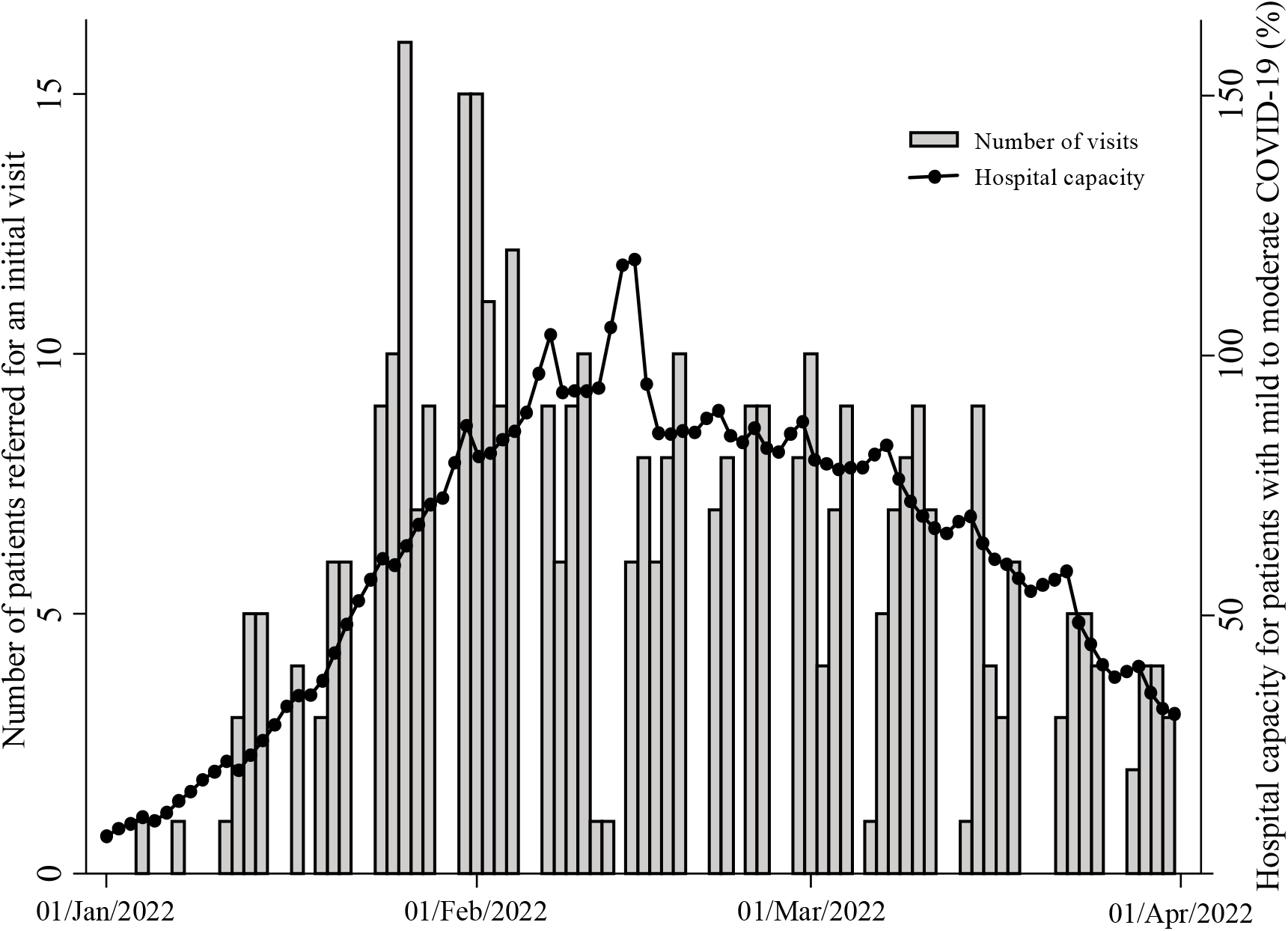
Number of visits upon the request from Osaka City Health Public Center and hospital capacity in Osaka. Note: The bars represents the number of doctor visits at home and the lines represents hospital capacity for patients with mild to moderate COVID-19 patients.We used the data on hospital capacity in Osaka was publicly available from https://www.pref.osaka.lg.jp/iryo/osakakansensho/corona_model.html (in Japanese). Abbreviations: COVID-19, Coronavirus disease

The results of the exploratory analysis are shown in Table 2. Obesity (Odds ratio (OR) 6.10 95% confidence interval (CI) 1.25 to 29.75), fever (OR 7.51, 95%CI 1.70 to 33.22), and gastrointestinal symptoms (OR 7.56, 95%C I1.57 to 36.40) were independently associated with an increased risk of receiving medical care beyond HaH care. Meanwhile, patients with a family physician (0R 0.06, 95%CI 0.008 to 0.44) and cough (OR 0.03, 0.003 to 0.38) were associated with a lower risk of receiving medical care beyond the HaH care.

**Table 2.** Factors associated with receiving medical care beyond the hospital-at-home care^*^ (n = 214)

| Characteristics | Odds ratio (95% confidence interval) | p-value |
| --- | --- | --- |
| **General** |  |  |
| Age | 1.01 (0.96 to 1.06) | 0.70 |
| Male gender | 1.21 (0.25 to 5.82) | 0.82 |
| Days from onset | 0.90 (0.72 to 1.13) | 0.37 |
| Unvaccinated<sup>†</sup> | 2.34 (0.57 to 9.60) | 0.24 |
| Ability to walk |  |  |
| Independent | reference |  |
| Minor help | 0.56 (0.005 to 154.13) | 0.95 |
| Major help | 5.57 (0.42 to 73.64) | 0.19 |
| Bedridden | 29.12 (0.001 to 685648.80) | 0.51 |
| Living with children under 18 | 0.18 (0.03 to 1.22) | 0.079 |
| Living alone | 0.36 (0.06 to 2.38) | 0.29 |
| Public assistance insurance | 0.94 (0.07 to 13.38) | 0.96 |
| Having a family physician | 0.06 (0.008 to 0.48) | 0.008 |
| Food procurement<sup>‡</sup> | 4.28 (0.21 to 85.43) | 0.34 |
| Weekend requests for HaH care<sup>§</sup> | 3.51 (0.67 to 18.44) | 0.14 |
| **Risk factors** |  |  |
| Chronic respiratory disease | 0.69 (0.11 to 4.45) | 0.70 |
| Hypertension | 2.04 (0.35 to 11.79) | 0.43 |
| Arrhythmia | 0.58 (0.02 to 13.89) | 0.73 |
| Obesity | 5.90 (1.19 to 29.11) | 0.029 |
| Hyperlipidemia | 0.56 (0.10 to 3.29) | 0.52 |
| Diabetes mellitus | 2.14 (0.28 to 6.20) | 0.46 |
| Chronic kidney disease | 14.64 (0.40 to 541.52) | 0.15 |
| Depression | 3.09 (0.15 to 64.30) | 0.47 |
| Dementia | 6.23 (0.02 to 1760.59) | 0.53 |
| Malignancy | 2.70 (0.24 to 30.02) | 0.42 |
| **Symptoms at the initial treatment** |  |  |
| Peripheral oxygen saturation |  |  |
| 96% ≤ | reference |  |
| 93% to 96% | 1.25 (0.13 to 12.44) | 0.85 |
| 93% ≥ | 5.19 (0.61 to 44.30) | 0.13 |
| Fever | 7.26 (1.62 to 32.46) | 0.009 |
| Cough | 0.03 (0.003 to 0.42) | 0.009 |
| Dyspnea | 1.99 (0.39 to 10.15) | 0.41 |
| Gastrointestinal symptom | 7.83 (1.59 to 38.55) | 0.011 |
| Water intake of less than 500ml per day | 3.52 (0.51 to 24.59) | 0.20 |
| Headache | 2.75 (0.57 to 13.13) | 0.20 |
| Sore throat | 3.07 (0.35 to 27.21) | 0.31 |
| Losing taste | 0.26 (0.04 to 1.69) | 0.16 |
| Fatigue | 0.69 (0.07 to 7.19) | 0.75 |
| Chest pain | 1.11 (0.24 to 5.19) | 0.90 |

Note: \* A composite outcome of any medical consultation outside of the HaH care team, hospitalization, or death due to any cause within 30 days from the initial treatment. †No vaccination or only first vaccination against Coronavirus disease. ‡Living with children under 18 years old. §Inability to procure food without stockpiles as declared by the individual. ¶Requests for HaH care made from Friday to Sunday. ||Only patients who needed doctor visits at home for the initial treatment were included, and the straight-line distance from the clinic where the initial treatment was provided to the patient's home was calculated using latitude and longitude. ¶Post-exertion oxygen saturation if applicable. Abbreviations: HaH, hospital at home

Table 3 shows the details of the treatment provided by the HaH care team. One-hundred twenty-nine (37%) patients were managed online alone without home visit, while 174 (50%) required only one home visit in addition to online treatment. The HaH team who visited patients requiring medical care beyond the HaH care more frequently administered intravenous fluids and prescribed corticosteroids.

**Table 3.** Details of the treatment provided by the hospital-at-home care team.

| Treatment | All (n = 348) | Received medical care solely by the HaH team (n = 311) | Received medical care beyond the HaH team<sup>†</sup> (n = 37) | *p*-value<sup>*</sup> |
| --- | --- | --- | --- | --- |
| Number of doctor visits at home |  |  |  | <0.001 |
| Online visit only | 129 (37) | 123 (40) | 6 (16) |  |
| 1 | 174 (50) | 155 (50) | 19 (51) |  |
| 2 | 40 (12) | 29 (9) | 11 (30) |  |
| 3 | 3 (1) | 3 (1) | 0 (0) |  |
| 4 or more | 2 (1) | 1 (0.3) | 1 (3) |  |
| Number of online doctor visits |  |  |  | 0.002 |
| In-home visit only | 50 (14) | 45 (15) | 5 (14) |  |
| 1 | 53 (15) | 42 (14) | 11 (30) |  |
| 2 | 72 (21) | 59 (19) | 13 (35) |  |
| 3 | 66 (19) | 64 (21) | 2 (5) |  |
| 4 or more | 107 (31) | 101 (33) | 6 (16) |  |
| Number of types of prescription drugs |  |  |  | 0.44 |
| None | 79 (23) | 68 (22) | 11 (30) |  |
| 1 to 3 | 137 (39) | 124 (40) | 13 (35) |  |
| 4 to 6 | 91 (26) | 80 (26) | 11 (30) |  |
| 7 or more | 41 (12) | 39 (13) | 2 (5) |  |
| Number of days of intravenous infusion |  |  |  | <0.001 |
| None | 316 (91) | 289 (93) | 27 (73) |  |
| 1 to 3 | 24 (7) | 18 (6) | 6 (16) |  |
| 4 or more | 8 (2) | 4 (1) | 4 (11) |  |
| Corticosteroids<sup>‡</sup> | 23 (7) | 17 (6) | 6 (16) | 0.013 |
| Remdesivir<sup>‡</sup> | 2 (1) | 1 (0.3) | 1 (3) | 0.070 |
| Molnupiravir<sup>‡</sup> | 9 (3) | 8 (3) | 1 (3) | 0.96 |
| Sotrovimab<sup>‡</sup> | 11 (3) | 10 (3) | 1 (3) | 0.87 |

Note: Values in parentheses shows percentage. Percentage calculations used a denominator that excluded missing values. <sup>\*</sup>*p*-values based on the Pearson's chi-squared test for categorical variables <sup>†</sup>A composite outcome of emergency room visits, hospitalization, or death due to any cause within 30 days from the initial treatment <sup>‡</sup>At least one dose of prescription of these drugs. Abbreviations: HaH, hospital at home

## Discussion

This study summarized the characteristics, treatments, and clinical outcomes of COVID-19 patients who received HaH care provided by a team of general and primary care clinics during the outbreak in Japan in early 2022. Our model allowed the avoidance of hospital visits for around 90% of patients who were not able to be triaged for emergency care through telephone consultation by the Health Center. Although 11% of the patients who were initially judged not to require hospitalization consequently received medical care beyond our team, the remaining patients were able to complete the treatment at home. No patients died during either HaH or hospitalization. We found several factors associated with an increased risk of receiving medical care beyond our team, including obesity, fever, and gastrointestinal symptoms, whereas having a physician and cough was associated with lower risk. Most patients required only one doctor visit or online visits.

Consistent with previous studies, our results showed that our HaH care model for COVID-19 patients is a useful alternative to alleviate the shortage of hospital staff and beds (7, 9-12). We further suggested that such models could be built by multiple clinics that normally provide primary care services. There were no deaths in the patients followed by the HaH care team, even though Osaka recorded the highest mortality in Japan during the period (13). Compared with the HaH care administered by only one clinic or hospital, our model is instantly applicable to many settings. It is difficult for a general clinic to be responsible for initial emergency HaH care over a long period. In our case, one of the 10 clinics was responsible for initial treatment on a rotating basis, with follow-up care provided by the clinic closest to the patient’s home. Of note, we did not intend that COVID-19 patients should insist on completing their treatment at home, especially when there is room in the hospital beds. However, during the COVID-19 outbreak, healthcare workers have suffered physical and psychological distress (17, 18). In such situations, it is important for the health of the entire community that primary care clinics support the hospital more than usual.

The factors associated with medical care beyond HaH care were partially inconsistent with previous reports (9, 19). Baseline oxygen saturation has been reported to be a prognostic factor for hospitalization during HaH care, but we could not confirm this finding (19). This may be due to the small sample size, or to the sampling method, i.e., only patients for whom the Health Center could not triage the need for hospitalization and who could not be treated urgently at a primary care clinic near their homes. In our sample, the majority of the patients admitted for hypoxia did not have an oxygen saturation below 93% at baseline. Studies revealed that COVID-19 symptoms were likely to worsen around day 7 (20, 21). Even in patients who were not hypoxic at baseline and were treated at home, the symptoms may worsen over time. Among known risk factors including unvaccinated status, obesity was the sole factor that was independently associated with worse outcome of our sample. This may be due to the small sample size, but obese patients with COVID-19 may be carefully monitored when received HaH care. Fever and gastrointestinal symptoms may increase the risk of medical care beyond the team, whereas having a family physician and cough was associated with a lower risk of worsening. Similar to oxygen saturation, our small sample size and sampling method may explain this result, and we could not find a reasonable explanation for this point.

Rotating initial treatment and online visits may reduce the burden on primary care providers in implementing the HaH care. In fact, nearly 40% of patients were able to be managed online alone, and nearly half required only one visit in addition to online treatment. Use of online consultation tools, such as video calls, could reduce the need for in-person treatment. A previous study showed that online physician visits were noninferior to physician visits at home for patients requiring acute care (22). Effective use of online medical care, with backups for in-home visits, may be key to the future expansion of HaH treatment for COVID-19.

Our study has several limitations. First, the present HaH model was designed as easy to construct, but as with previous reports, it was limited in its generalizability. Our sample included only patients for whom the health center could not triage the need for hospitalization, and no primary care clinic near their homes could treat their emergencies. However, during an outbreak, such patients would likely strain the hospital emergency department, and should be handled at the primary care level. Additionally, the median straight-line distance from the clinic responsible for the initial treatment to patients’ home was around 5.7 km in the present study. Further studies are needed to assess the usefulness of HaH care in rural areas. Second, due to the small sample size and lack of statistical power, it is uncertain if known risk factors were not associated with poor outcomes in this population. The findings of our exploratory analysis should be interpreted with cautions as we examined many factors, and could not explain some of the mechanisms. Third, policies for emerging infectious diseases such as COVID-19 can change dramatically with time. During the study period, patients with COVID-19 had to be quarantined for 10 days, but this has now been reduced to 7 days (23). As policies change, people’s behavior also changes (24, 25). The usefulness of HaH care for COVID-19 would therefore need to be tested repeatedly.

Despite the limitations above, this study is the first report of HaH care provided by a team of local primary care clinics. In accordance with the rigorous methodology, including the pre-specified protocol and adherence to the standard reporting guideline (16), we described the characteristics, treatment, and clinical outcomes of patients who received HaH care in detail.

## Conclusion

In conclusion, the HaH care model with a team of primary care clinics was able to triage patients with COVID-19 who required immediate hospitalization without going through a hospital, and most of the remaining patients were treated at home. The instant model may lighten the burden on hospitals during the COVID-19 outbreak. The effective use of online physician visits may also play an important role. Although our HaH care resulted in no deaths among patients followed up at home, more research is needed to predict patients who will not be successfully treated by HaH care alone.

## Data Availability

The data are available to academic researchers upon a reasonable request. 

## Footnotes

## Acknowledgement

We thank all the participating research sites and the time and support from KISA2-Tai members including medical doctors, nurses, physical therapists, pharmacists, medical assistants, care workers, and local collaborators. We thank Ms. Chiharu Tojo for helping the data extraction from the electric records. We are also grateful for the support from Ms. Chihiro Miyagawa, Ms. Mitsuko Shinohara, and N-STAFF. Co., Ltd who assisted in day-to-day management of the KISA2-Tai activities.

## Contributors

YTs had full access to all data used in the study and takes responsibility for both the integrity of the data and the accuracy of the data analysis. All authors developed the study’s concept and design. YTs, MK, TOk, SY, MT, NM, MN, YTe, TT, and YK acquired the data. YTs, TOg, and YMi analyzed the data. All authors interpreted the data. YTs, TOg, and YMi drafted the manuscript. MK, Tok, SY, MT, YTe, TT, TT, YK, and TMo critically revised the manuscript for important intellectual content. All the authors provided their final approval of the version submitted for publication and agreed that they were accountable for all aspects of this study.

## Data sharing

The data are available to academic researchers upon a reasonable request.

