## Supplementary files for "A novel hospital-at-home model for patients with COVID-19 built by a team of local primary care clinics and clinical outcomes: A multi-center retrospective cohort study"

Text S1. The KISA2-Tai collaborators

Table S1. The STROBE checklist

Table S2. Difference between the protocol and publication

Table S3. Characteristics of patients who needed immediate hospitalization after the first treatment by the hospital at home care team

Table S4. The number of missing values in each variable

Table S5. Summary of the reasons for the requirement of medical care beyond the hospital-at-home team

**Text S1. The KISA2-Tai collaborators**

Ui Nakagawa, Oku Medical Clinic, Osaka

Keisuke Shirosaka, Oku Medical Clinic, Osaka

Nobuyuki Kajiwara, Oku Medical Clinic, Osaka

Yuko Makuuchi, Kassai Medical Clinic, Osaka

Aya Shibata, Kassai Medical Clinic, Osaka

Masaru Mito, General Incorporated Association KISA2-Tai, Osaka

Anna Suzuki, General Incorporated Association KISA2-Tai, Osaka

Hiroki Tagomori, General Incorporated Association KISA2-Tai, Osaka

**Table S1. The STROBE checklist**

|  | Item No | Recommendation | Pages |
| --- | --- | --- | --- |
| **Title and abstract** | 1 | (*a*) Indicate the study’s design with a commonly used term in the title or the abstract | 1 |
|  |  | (*b*) Provide in the abstract an informative and balanced summary of what was done and what was found | 2 |
| Introduction | | |  |
| Background/rationale | 2 | Explain the scientific background and rationale for the investigation being reported | 4 |
| Objectives | 3 | State specific objectives, including any prespecified hypotheses | 4 |
| Methods | | |  |
| Study design | 4 | Present key elements of study design early in the paper | 5 |
| Setting | 5 | Describe the setting, locations, and relevant dates, including periods of recruitment, exposure, follow-up, and data collection | 5 |
| Participants | 6 | (*a*) Give the eligibility criteria, and the sources and methods of selection of participants. Describe methods of follow-up | 5 |
|  |  | (*b*) For matched studies, give matching criteria and number of exposed and unexposed | NA |
| Variables | 7 | Clearly define all outcomes, exposures, predictors, potential confounders, and effect modifiers. Give diagnostic criteria, if applicable | 6 |
| Data sources/ measurement | 8* | For each variable of interest, give sources of data and details of methods of assessment (measurement). Describe comparability of assessment methods if there is more than one group | *6* |
| Bias | 9 | Describe any efforts to address potential sources of bias | 7 |
| Study size | 10 | Explain how the study size was arrived at | 7 |
| Quantitative variables | 11 | Explain how quantitative variables were handled in the analyses. If applicable, describe which groupings were chosen and why | 6 |
| Statistical methods | 12 | (*a*) Describe all statistical methods, including those used to control for confounding | 7 |
|  |  | (*b*) Describe any methods used to examine subgroups and interactions | 7 |
|  |  | (*c*) Explain how missing data were addressed | 7 |
|  |  | (*d*) If applicable, explain how loss to follow-up was addressed | 7 |
|  |  | (*e*) Describe any sensitivity analyses | Supplement |
| Results | | |  |
| Participants | 13* | (a) Report numbers of individuals at each stage of study—eg numbers potentially eligible, examined for eligibility, confirmed eligible, included in the study, completing follow-up, and analysed | 8 |
|  |  | (b) Give reasons for non-participation at each stage | 8 |
|  |  | (c) Consider use of a flow diagram | Figure 1 |
| Descriptive data | 14* | (a) Give characteristics of study participants (eg demographic, clinical, social) and information on exposures and potential confounders | 8, Table 1 |
|  |  | (b) Indicate number of participants with missing data for each variable of interest | Supplement |
|  |  | (c) Summarise follow-up time (eg, average and total amount) | 8 |
| Outcome data | 15* | Report numbers of outcome events or summary measures over time | 8, Table 1 |
| Main results | 16 | (*a*) Give unadjusted estimates and, if applicable, confounder-adjusted estimates and their precision (eg, 95% confidence interval). Make clear which confounders were adjusted for and why they were included | Table 2 |
|  |  | (*b*) Report category boundaries when continuous variables were categorized | 8, Table 1, Table 2 |
|  |  | (*c*) If relevant, consider translating estimates of relative risk into absolute risk for a meaningful time period | NA |
| Other analyses | 17 | Report other analyses done—eg analyses of subgroups and interactions, and sensitivity analyses | Supplement |
| Discussion | | |  |
| Key results | 18 | Summarise key results with reference to study objectives | 9 |
| Limitations | 19 | Discuss limitations of the study, taking into account sources of potential bias or imprecision. Discuss both direction and magnitude of any potential bias | 10 |
| Interpretation | 20 | Give a cautious overall interpretation of results considering objectives, limitations, multiplicity of analyses, results from similar studies, and other relevant evidence | 9 |
| Generalisability | 21 | Discuss the generalisability (external validity) of the study results | 10 |
| Other information | | |  |
| Funding | 22 | Give the source of funding and the role of the funders for the present study and, if applicable, for the original study on which the present article is based | 11 |

**Supplementary Table 2. Difference between the protocol and final publication.**

| **Item** | **Initial plan in the protocol and reason for the amendments.** |
| --- | --- |
| 1 | We could not obtain the data of medical cost, which is one of the pre-specified secondary outcomes. |
| 2 | Although not originally planned, we contacted the patients if a primary endpoint was missing. |
| 3. | We added the narrative summary of the reason for the medical care beyond the hospital from the electric records. |
| 4. | We planned a pre-specified sensitivity analysis focusing on the period under a declared state of emergency, however, we could not perform it due to the small sample size. |

| **Table S3. Characteristics of patients who needed immediate hospitalization^*^ after the first triage by the hospital-at-home care team (n = 34)** | |
| --- | --- |
| **Characteristics** |  |
| ***General*** |  |
| Age | 76.5 (53 to 84) |
| Male gender | 22 (65) |
| Days from onset | 7 (5 to 9) |
| Unvaccinated^†^ | 14 (45) |
| Ability to walk |  |
| Independent | 22 (71) |
| Major help | 6 (19) |
| Bedridden | 3 (10) |
| Living with young children | 7 (23) |
| Living alone | 9 (27) |
| Public assistance insurance | 5 (15) |
| Having a family physician | 24 (80) |
| Food procurement^‡^ | 28 (90) |
| Weekend requests for HaH care^§^ | 9 (26) |
| Distance (km) ^\|\|^ | 5.3 (3.2 to 9.4) |
| ***Symptoms at the initial treatment*** |  |
| Chronic respiratory disease | 6 (18) |
| Hypertension | 10 (29) |
| Arrhythmia | 2 (6) |
| Obesity | 8 (24) |
| Hyperlipidemia | 4 (12) |
| Diabetes mellitus | 5 (15) |
| Chronic kidney disease | 0 (0) |
| Depression | 1 (3) |
| Dementia | 5 (15) |
| Malignancy | 5 (15) |
| Pregnant | 1 (3) |
| Fever | 12 (35) |
| Peripheral oxygen saturation^¶^ |  |
| 96 ≤ | 15 (44) |
| 93 to 96 | 1 (3) |
| 93 ≥ | 18 (53) |
| Cough | 22 (71) |
| Dyspnea | 13 (42) |
| Gastrointestinal symptom | 10 (32) |
| Water intake of less than 500ml per day | 7 (23) |
| Headache | 6 (20) |
| Sore throat | 10 (33) |
| Losing taste | 8 (29) |
| Fatigue | 27 (87) |
| Chest pain | 7 (23) |
| Note: Values in parentheses shows percentage or interquartile range. Percentage calculations used a denominator that excluded missing values. ^*^Patients who were hospitalized after the first doctor visit at home or online visit. ^†^A composite outcome of emergency room visits, hospitalization, or death due to any cause within 30 days from the initial treatment. ^‡^No vaccination or only first vaccination against Coronavirus disease ^§^Living with children under 18 years old. ^§^Inability to procure food without stockpiles as declared by the individual. ^\|\|^Requests for HaH care made from Friday to Sunday. ^¶^Only patients who needed doctor visits at home for the initial treatment were included (n = 30), and the straight-line distance from the clinic where the initial treatment was provided to the patient's home was calculated using latitude and longitude. **^**^**Post-exertion oxygen saturation if applicable. Abbreviations: HaH, hospital at home | |

| **Table S4. Number of missing values in the variables** | | | |
| --- | --- | --- | --- |
| **Characteristics** | **All (n =348)** | **Received medical care solely by the HaH care (n = 311)** | **Received medical care beyond the HaH care^*^ (n = 37)** |
| ***General*** |  |  |  |
| Age | 0 | 0 | 0 |
| Male gender | 0 | 0 | 0 |
| Day from onset | 17 | 17 | 0 |
| Unvaccinated^†^ | 27 (8) | 25 (8) | 2 (5) |
| Ability to walk | 29 (8) | 25 (8) | 4 (11) |
| Living with children under 18 | 48 (14) | 42 (14) | 5 (14) |
| Living alone | 31 (9) | 27 (9) | 4 (11) |
| Public assistance insurance | 26 (7) | 24 (8) | 2 (5) |
| Having a family physician | 57 (16) | 55 (18) | 2 (5) |
| Food procurement^‡^ | 55 (16) | 51 (16) | 4 (11) |
| Weekend requests for HaH care^§^ | 0 | 0 | 0 |
| Distance (km) ^\|\|^ | 0 | 0 | 0 |
| ***Risk factors*** |  |  |  |
| Chronic respiratory disease | 0 | 0 | 0 |
| Hypertension | 0 | 0 | 0 |
| Arrhythmia | 0 | 0 | 0 |
| Obesity | 0 | 0 | 0 |
| Hyperlipidemia | 0 | 0 | 0 |
| Diabetes mellitus | 0 | 0 | 0 |
| Chronic kidney disease | 0 | 0 | 0 |
| Depression | 0 | 0 | 0 |
| Dementia | 0 | 0 | 0 |
| Malignancy | 0 | 0 | 0 |
| Pregnant | 0 | 0 | 0 |
| ***Symptoms at the initial treatment*** |  |  |  |
| Peripheral oxygen saturation^¶^ | 26 (7) | 20 (6) | 5 (14) |
| Fever | 0 | 0 | 0 |
| Cough | 19 (5) | 18 (6) | 1 (3) |
| Dyspnea | 18 (5) | 18 (6) | 0 |
| Gastrointestinal symptom | 24 (7) | 22 (7) | 2 (5) |
| Water intake of less than 500ml per day | 23 (7) | 21 (9) | 2 (5) |
| Headache | 30 (9) | 29 (9) | 1 (3) |
| Sore throat | 31 (9) | 29 (9) | 2 (5) |
| Losing taste | 41 (12) | 37 (12) | 3 (8) |
| Fatigue | 23 (7) | 21 (7) | 1 (3) |
| Chest pain | 32 (9) | 31 (10) | 1 (3) |
| ***Treatments*** |  |  |  |
| Number of doctor visits at home | 0 | 0 | 0 |
| Number of online doctor visits | 0 | 0 | 0 |
| Number of types of prescription drugs | 0 | 0 | 0 |
| Number of days of intravenous infusion | 0 | 0 | 0 |
| Corticosteroids^‡^ | 0 | 0 | 0 |
| Remdesivir^‡^ | 0 | 0 | 0 |
| Molnupiravir^‡^ | 0 | 0 | 0 |
| Sotrovimab^‡^ | 0 | 0 | 0 |

Note: Values in parentheses shows percentage. Percentage calculations used a denominator that excluded missing values. ^*^A composite outcome of any medical consultation outside of the HaH care team, hospitalization, or death due to any cause within 30 days from the initial treatment. ^†^No vaccination or only first vaccination against Coronavirus disease. ^‡^Living with children under 18 years old. ^§^Inability to procure food without stockpiles as declared by the individual. ^§^Requests for HaH care made from Friday to Sunday. ^||^Only patients who needed doctor visits at home for the initial treatment were included, and the straight-line distance from the clinic where the initial treatment was provided to the patient's home was calculated using latitude and longitude. ^¶^Post-exertion oxygen saturation if applicable. Abbreviations: HaH, hospital-at-home

| **Table S5. The reasons for the requirement of the medical care beyond the hospital-at-home care (n = 32)** | | | | | |
| --- | --- | --- | --- | --- | --- |
| **Age** | **Gender** | **Oxygen saturation at baseline** | **Days from onset to outcome** | **Types of outcomes** | **Reason for the medical care beyond the HaH care** |
| 60-65 | Male | 98 | 12 | Discharged alive after hospitalization | Abdominal pain suspicious for appendicitis |
| 0-5 | Female | missing | 7 | Medical consultation other than hospitalization | Acute exacerbation of asthma |
| 60-65 | Male | 93 | 15 | Discharged alive after hospitalization | Acute exacerbation of heart failure |
| 45-50 | Female | 98 | 11 | Discharged alive after hospitalization | Bowel obstruction required a surgery |
| 50-55 | Female | 99 | 11 | Discharged alive after hospitalization | Diabetic ketoacidosis |
| 60-65 | Female | missing | 11 | Discharged alive after hospitalization | Drug allergy |
| 45-50 | Male | 99 | 7 | Medical consultation other than hospitalization | Family physician took over the treatment |
| 70-75 | Male | 96 | 2 | Medical consultation other than hospitalization | Family physician took over the treatment |
| 70-75 | Female | 96 | 13 | Medical consultation other than hospitalization | Family physician took over the treatment |
| 0-5 | Male | 99 | 8 | Medical consultation other than hospitalization | Family physician took over the treatment |
| 50-55 | Female | 99 | 4 | Discharged alive after hospitalization | Hemodialysis patients |
| 45-50 | Female | 96 | 5 | Discharged alive after hospitalization | Hypoxia |
| 45-50 | Female | 98 | 10 | Discharged alive after hospitalization | Hypoxia |
| 50-54 | Male | 95 | 9 | Discharged alive after hospitalization | Hypoxia |
| 90-95 | Male | 96 | 7 | Discharged alive after hospitalization | Hypoxia and unclear consciousness |
| 45-50 | Female | 99 | 15 | Discharged alive after hospitalization | Impaired consciousness with hypokalemia |
| 80-85 | Male | 98 | 11 | Discharged alive after hospitalization | Persistent appetite loss suspicious for diseases other than COVID-19 |
| 75-80 | Female | 96 | 12 | Discharged alive after hospitalization | Persistent appetite loss suspicious for diseases other than COVID-19 |
| 50-55 | Female | 98 | 14 | Discharged alive after hospitalization | Persistent fatigue with hyponatremia |
| 70-75 | Male | 95 | 13 | Discharged alive after hospitalization | Persistent fever |
| 75-80 | Male | 95 | 17 | Discharged alive after hospitalization | Persistent fever |
| 60-65 | Male | 96 | 10 | Discharged alive after hospitalization | Persistent fever |
| 50-55 | Male | 95 | 13 | Discharged alive after hospitalization | Persistent fever with immunosuppressive status |
| 85-90 | Female | 96 | 7 | Discharged alive after hospitalization | Persistent fever with unstable heart failure |
| 65-70 | Male | 93 | 10 | Discharged alive after hospitalization | Persistent hypoxia |
| 45-50 | Male | 94 | 13 | Discharged alive after hospitalization | Persistent hypoxia and fever |
| 20-25 | Female | 99 | 6 | Discharged alive after hospitalization | Persistent hypoxia and fever |
| 80-85 | Male | 93 | 5 | Discharged alive after hospitalization | Persistent hypoxia and fever |
| 55-60 | Male | 97 | 12 | Discharged alive after hospitalization | Persistent hypoxia and fever |
| 75-80 | Male | 86 | 7 | Discharged alive after hospitalization | Persistent hypoxia and fever |
| 55-60 | Male | 88 | 16 | Discharged alive after hospitalization | Persistent hypoxia and fever |
| 45-50 | Female | 94 | 12 | Discharged alive after hospitalization | Persistent hypoxia and fatigue despite a 10-day treatment |
| 90-95 | Female | 97 | 16 | Discharged alive after hospitalization | Persistent hypoxia suspicious for aspiration pneumonia |
| 65-70 | Female | 96 | 6 | Discharged alive after hospitalization | Severe dehydration in addition to an end-stage cancer |
| 95-100 | Female | 98 | 10 | Medical consultation other than hospitalization | Suspicious for bacterial pneumonia |
| 15-20 | Male | 99 | 11 | Medical consultation other than hospitalization | The symptoms became mild, and the patient wished to be followed up at the hotel |
| 60-65 | Male | 97 | 8 | Medical consultation other than hospitalization | The symptoms became mild, and the patient wished to be followed up at the hotel |
